# Performance study of a point-of-care antigen test during the SARS-CoV-2 Delta to Omicron variant transition in the USA

**DOI:** 10.1101/2022.05.11.22274962

**Authors:** Paul K Drain, Gregory Chiklis, Poppy Guest, Nigel M Lindner, Jayne E Ellis

**Affiliations:** University of Washington, Seattle, WA, USA; Medical Research Network, Franklin, MA, USA; LumiraDx Ltd, London, UK

**Author notes:** Corresponding author: Jayne E Ellis, PhD.

**Keywords:** Antigen test, Delta variant, LumiraDx, Omicron variant, SARS-CoV-2

## Abstract

**Introduction:** Concerns have been raised regarding the accuracy of diagnostic antigen testing for the severe acute respiratory syndrome coronavirus 2 (SARS-CoV-2) Omicron variant. We compared the performance of the LumiraDx SARS-CoV-2 Antigen Test between symptomatic participants recruited prospectively during the Delta to Omicron variant transition in the USA.

**Methods:** Two paired anterior nasal swabs were collected from each participant (adults and children) within 12 days of symptom onset between November 24^th^, 2021 and February 1^st^, 2022, during which time Omicron replaced Delta as the dominant variant in the sample population. Swabs were tested by the LumiraDx SARS-CoV-2 Antigen Test and compared using real-time polymerase chain reaction (RT-PCR) reference testing. Reference samples identified as positive were sequenced to identify the SARS-CoV-2 variant. Positive percent agreement (PPA) was calculated, with results stratified by RT-PCR cycle threshold (Ct).

**Results:** Of the 38 participants for whom LumiraDx SARS-CoV-2 Antigen Test results were available, 36 were confirmed positive by RT-PCR. Overall, PPA of the LumiraDx SARS-CoV-2 Antigen Test was 94.7% (95% confidence interval: 82.3%, 99.4%) and PPA was 100% for samples with a Ct <33. Sufficient viral load for sequencing was present in nine samples (six Delta, three Omicron), all of which returned a positive result using the LumiraDx SARS-CoV-2 Antigen Test. There were no performance differences observed between participants with the Delta and Omicron variants.

**Conclusions:** SARS-CoV-2 differences between Delta and Omicron variant mutations did not affect the performance of the LumiraDx SARS-CoV-2 Antigen Test which detects the nucleocapsid protein antigen. The LumiraDx SARS-CoV-2 Antigen Test can be a useful antigen test to diagnose emerging variants of coronavirus disease 2019.

## Introduction

On November 26^th^, 2021, the World Health Organization (WHO) added another variant of severe acute respiratory syndrome coronavirus 2 (SARS-CoV-2), named Omicron, to the list of variants of concern (VOC) [1]. Since the Omicron variant has 34 VOC mutations in the spike (S) protein, there was concern regarding increased transmissibility and immune escape as cases were rising in people who were either vaccinated or had experienced previous SARS-CoV-2 infections [2]. Within weeks, the Omicron variant rapidly spread throughout South Africa and was subsequently identified in 50 countries around the world [1, 3, 4]. The evidence suggested that a full vaccination course was inadequate to protect against serious disease complications, leading to booster vaccination programs being accelerated and expanded [5]. Studies investigating the severity of coronavirus disease 2019 (COVID-19) during the Omicron outbreak reported decreased severity, resulting in fewer patients hospitalized compared with during the peak of the Delta outbreak [6] (Preprint: Wang L, et al. doi: https://doi.org/10.1101/2021.12.30.21268495). However, healthcare systems could still be put under heavy strain as the unprecedented transmissibility of the Omicron variant could cause a dramatic increase in staff self-isolation and absences [7].

Accurate and timely testing of both symptomatic and asymptomatic individuals is essential for containment of the rapid spread of this SARS-CoV-2 variant [8]. There are concerns that diagnostic antigen tests are less able to detect the Omicron variant due to the many mutations present in all structural proteins compared with previous strains of the virus. Preliminary results from the US National Institutes of Health RADx program, which studied the performance of antigen tests using patient samples containing the Omicron variant, suggested that these tests detect the Omicron variant but may have reduced sensitivity [9]. Other studies have also reported reduced sensitivity for the Omicron variant with some antigen tests [10, 11]. The reduced sensitivity reported did not appear to be due to reduced viral load in samples,[11] and a study of different sampling methods noted that nasal swab samples had higher sensitivity compared with oral (cheek or throat) swabs, suggesting that this common sampling method is the most effective for detecting the Omicron variant [12].

A further in-depth evaluation using patient samples with live virus is necessary to understand the extent of any potential reduced sensitivity, in order to make informed decisions when implementing a SARS-CoV-2 antigen test strategy. The LumiraDx SARS-CoV-2 Antigen Test has been designed with antibodies that are raised to capture nucleocapsid (N) proteins in SARS-CoV-2-positive samples. The choice of capturing the N protein over the S protein was made because the N protein is more abundantly present, with fewer reported mutations in common variants [13, 14]. This interim study analysis investigated the performance of the LumiraDx SARS-CoV-2 Antigen Test in samples of patients infected with the SARS-CoV-2 Delta and Omicron variants compared using real-time polymerase chain reaction (RT-PCR) reference testing. During the period of testing, Omicron replaced Delta as the dominant SARS-CoV-2 variant in the state of Massachusetts, USA,[15] which allowed for evaluation of the LumiraDx SARS-CoV-2 Antigen Test performance in a head-to-head comparison of the two variants in real time.

## Methods

### Study design

Prospectively collected samples were tested using the LumiraDx SARS-CoV-2 Antigen Test at one site in Franklin (MA, USA) from November 24^th^, 2021 to February 1^st^, 2022. Anterior nasal swab samples were collected from adults and children who presented with symptoms indicative of SARS-CoV-2 infection. Eligible participants were those who presented with symptoms of SARS-CoV-2 infection within 12 days of symptom onset. Presenting symptoms were defined as fever, cough, shortness of breath, difficulty breathing, tiredness, body aches, runny nose, sore throat, loss of smell, loss of taste, and vomiting. This study has been conducted in accordance with the Helsinki Declaration. The study received ethical approval from the Diagnostics Investigational Review Board (IRB project #1021-14). All participants, or parents or guardians of minor participants, provided informed consent.

Two paired anterior nasal swabs (Nasal FLOQ Swab 502CS01; Copan Diagnostics Inc; Murrieta, CA, USA) were collected from each participant by inserting a swab in each nostril and then exchanging the swab into the second nostril to ensure that a sample from each nostril was collected on each swab, and to minimize bias between swabs. One swab was then placed into a proprietary extraction buffer for the LumiraDx SARS-CoV-2 Antigen Test, and the other swab was placed into 3 mL of viral transport medium (VTM; Universal Viral Transport System; Becton, Dickinson and Company; Franklin Lakes, NJ, USA). The swabs in VTM were tested fresh and according to the manufacturer’s instructions by RT-PCR using Xpert Xpress SARS-CoV-2/Flu/RSV (Cepheid, Sunnyvale, CA, USA). Reference VTM was then sent for sequencing of SARS-CoV-2 ribonucleic acid at Wisconsin Diagnostic Laboratories (Milwaukee, WI, USA).

### Statistical analysis

Positive percent agreement (PPA) with RT-PCR was assessed, with 95% confidence interval (CI) calculated using the Wilson Score method. A further evaluation was performed using days per symptom onset and cycle threshold (Ct) values. To highlight the change in SARS-CoV-2 variant dominance, prevalence data for the region of sample collection (Franklin, MA, USA) was obtained from the website: https://covariants.org/ [15]. This data was used to plot against the results from the LumiraDx Antigen Test, over the testing period, according to RT-PCR Ct value.

## Results

Overall, 93 participants were recruited to the study. Of the 38 eligible SARS-CoV-2 positive participants with confirmed by RT-PCR, the majority (28, 74%) were aged 22 to 59 years, and more than half were female (20, 53%). RT-PCR Ct values in the positive population ranged from 15 to 40, with the majority under 30 (35, 92%). All participants were tested between 1 and 7 days following symptom onset (**Figure 1**). Overall, 36 of the 38 participant samples tested positive using the LumiraDx SARS-CoV-2 Antigen Test, resulting in a PPA of 94.7% (95% CI: 82.3, 99.4). This increased to 100% (95% CI: 90.4, 100) if the two samples with a Ct >33 were excluded from the analysis. The two false negative (FN) samples came from patients with recent symptom onset, and showed high Ct values indicative of low viral load (**Table 1, Figure 1**).

**Table 1:** Performance of the LumiraDx SARS-CoV-2 Antigen Test based on TFSO DFSO, days from symptom onset; FN, false negative; POS, positive; PPA, positive percent agreement; SARS-CoV-2, severe acute respiratory syndrome coronavirus 2; TP, true positive

| ≤DFS0 | TP | FN | POS | PPA |
| --- | --- | --- | --- | --- |
| 0 | 0 | 0 | 0 | NA |
| 1 | 6 | 1 | 7 | 85.7% |
| 2 | 17 | 1 | 18 | 94.4% |
| 3 | 20 | 2 | 22 | 90.9% |
| 4 | 29 | 2 | 31 | 93.5% |
| 5 | 33 | 2 | 35 | 94.3% |
| 6 | 34 | 2 | 36 | 94.4% |
| 7 | 36 | 2 | 38 | 94.7% |
| 8 | 36 | 2 | 38 | 94.7% |
| 9 | 36 | 2 | 38 | 94.7% |
| 10 | 36 | 2 | 38 | 94.7% |
| 11 | 36 | 2 | 38 | 94.7% |
| 12 | 36 | 2 | 38 | 94.7% |

**Figure 1:**
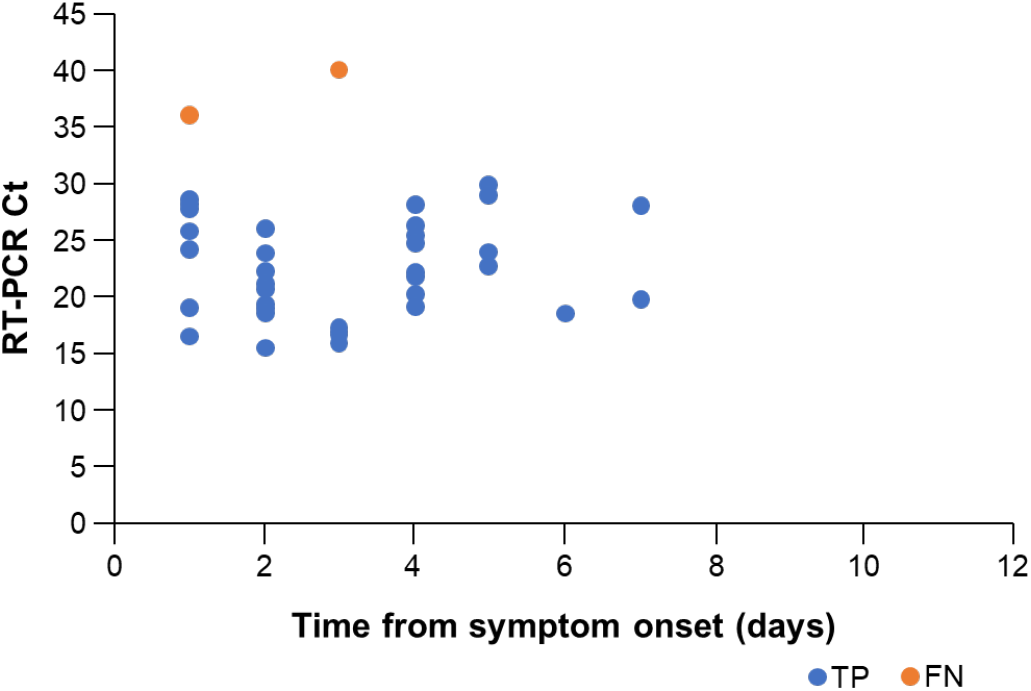
Performance of the LumiraDx SARS-CoV-2 Antigen Test based on Ct subsets and time from symptom onset Ct, cycle threshold; FN, false negative; RT-PCR, real-time polymerase chain reaction; SARS-CoV-2, severe acute respiratory syndrome coronavirus 2; TP, true positive

Of the samples collected in this study, nine (23%) were found to have sufficient viral load for sequencing. Of these, six samples collected between December 3^rd^ and 14^th^, 2021 were found to contain Delta variants, and three samples, collected on December 22^nd^, 2021 contained the Omicron variant. All nine samples tested positive for SARS-CoV-2 by the LumiraDx SARS-CoV-2 Antigen Test. Overall, the performance of the LumiraDx SARS-CoV-2 Antigen Test compared with RT-PCR was consistent across the period of the study, despite the change in the dominant variant in the population over that time (**Figure 2**).

**Figure 2:**
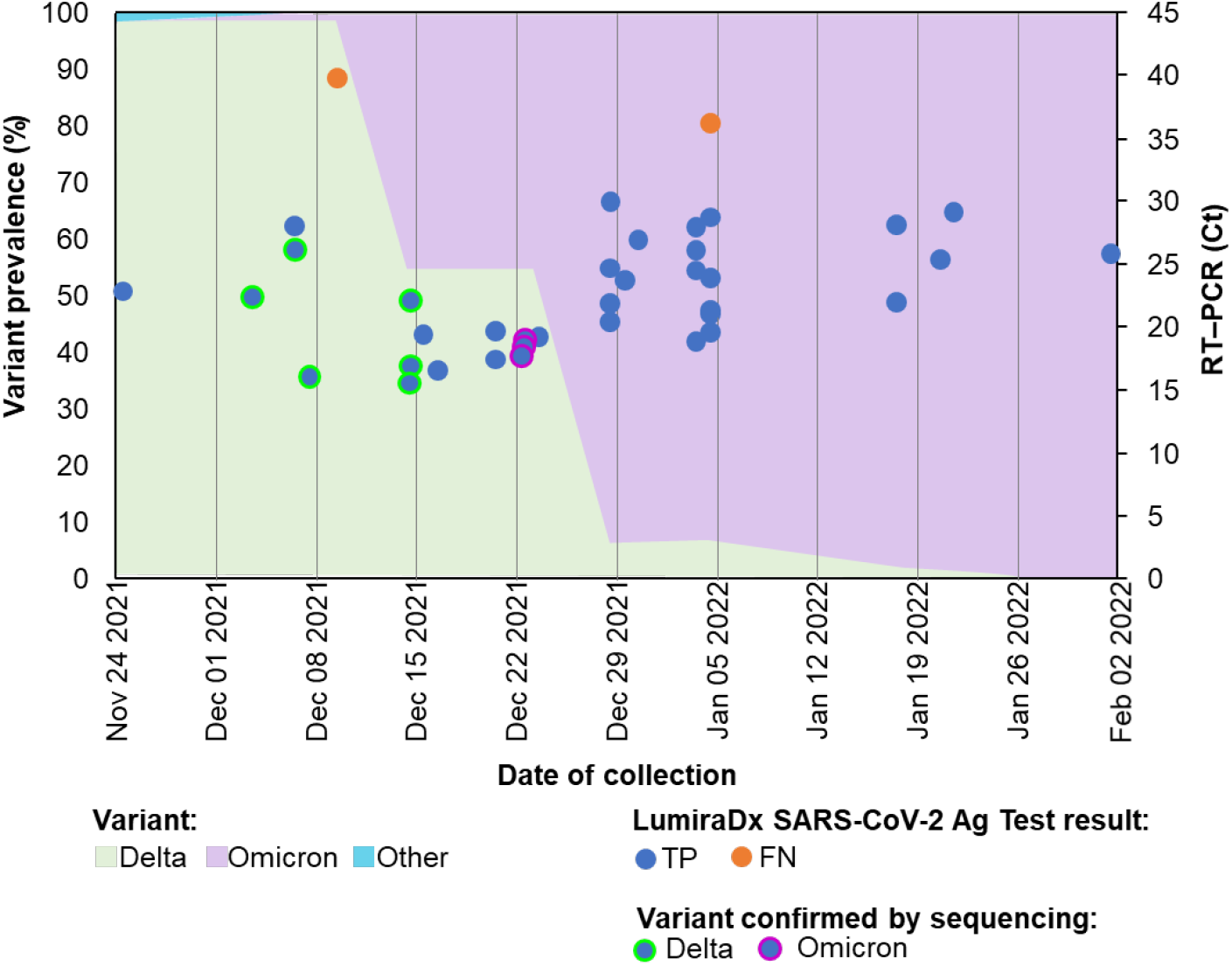
Results from LumiraDx Antigen Test over the testing period according to RT-PCR Ct value, plotted against variant sequencing data for the region of sample collection (Franklin, MA, USA) [15] Ag, antigen; Ct, cycle threshold; FN, false negative; RT-PCR, real-time polymerase chain reaction; SARS-CoV-2, severe acute respiratory syndrome coronavirus 2; TP, true positive

## Discussion

Since the WHO added Omicron to the list of VOCs, concerns have been raised about the effectiveness of antigen tests for the diagnosis of COVID-19 owing to the many mutations it contains [1, 9]. Results of this study demonstrate that the LumiraDx SARS-CoV-2 Antigen Test is able to detect the presence of the SARS-CoV-2 Omicron variant in RT-PCR-confirmed patient samples. Furthermore, the study data indicates that the performance of the LumiraDx SARS-CoV-2 Antigen Test was comparable to performance previously reported in symptomatic (97.6% sensitivity) and asymptomatic (82.1% PPA) individuals [16, 17]. Although only nine of the samples collected in this study population could be sequenced, the LumiraDx SARS-CoV-2 Antigen Test correctly identified all nine positive participants, irrespective of the SARS-CoV-2 variant of the infection. The timing of the samples with known variants (six Delta, three Omicron) was also consistent with state-level sequencing data, suggesting the variance prevalence in the study population matched what was reported for the wider state of Massachusetts [15]. Therefore, as the overall performance of the LumiraDx SARS-CoV-2 Antigen Test compared to RT-PCR was consistent throughout the study period, correctly identifying all RT-PCR positive cases with Ct <33, it can be inferred that the LumiraDx SARS-CoV-2 Antigen Test performance was not affected by the switch in the dominant SARS-CoV-2 variant from Delta to Omicron. Interestingly, the two samples reported as FN were obtained soon after the onset of symptoms and exhibited low viral loads, based on the high Ct values. This may be consistent with the milder infection seen with the Omicron variant. The results of this study are in line with a recent study in which the limit of detection (LoD) of the LumiraDx SARS-CoV-2 Antigen Test was determined for the Omicron variant in comparison with the WA1 strain, using live virus. The authors found a LoD of 2.5×10^2^ PFU/mL for both Omicron and WA1, indicating that the LoD for the LumiraDx SARS-CoV-2 Antigen Test was at least as good for the Omicron as for the WA1 strain (Preprint: Stanley S, et al. doi: https://doi.org/10.1101/2022.01.28.22269968).

A limitation of this study was that not all of the collected samples could be sequenced, due to a minimal viral load requirement for sequencing; samples with a Ct>28 could not be sequenced. Of the collected samples, only 23% contained sufficient sample for sequencing. However, during the period of testing, Omicron replaced Delta as the dominant SARS-CoV-2 variant in the state of Massachusetts[15]. State-wide, Omicron accounted for only 1% of sequenced SARS-CoV-2 samples in the week commencing November 29^th^, 2021. However, this increased to 99% of sequenced cases in the week commencing January 10^th^, 2022 [15]. Figure 2 highlights the shift in SARS-CoV-2 variant dominance from Delta to Omicron over time, indicating that the tested samples were a mixture of the two variants (**Figure 2**), as confirmed by the sequenced samples.

## Conclusion

The performance of the test was not affected by the mutations of the Omicron variant [16, 17]. The LumiraDx SARS-CoV-2 Antigen Test can effectively be used to diagnose SARS-CoV-2 and help to contain the spread of the virus.

## Data Availability

All data produced in the present study are available upon reasonable request to the author

## Acknowledgments

The study was funded by LumiraDx. The authors acknowledge Viola Kooij, PhD, of imc (integrated medhealth communication), for medical writing support.

## References

1. World Health Organisation. Classification of Omicron (B.1.1.529): SARS-CoV-2 Variant of Concern. Available from: https://www.who.int/news/item/26-11-2021-classification-of-omicron-(b.1.1.529)-sars-cov-2-variant-of-concern.

2. European Centre for Disease Prevention and Control. SARS-CoV-2 variants of concern as of 13 January 2022. 2022. Available from: https://www.ecdc.europa.eu/en/covid-19/variants-concern.

3. Meo SA, Meo AS, Al-Jassir FF, Klonoff DC. Omicron SARS-CoV-2 new variant: global prevalence and biological and clinical characteristics. Eur Rev Med Pharmacol Sci. 2021;25:8012–8.

4. Abdullah F. Tshwane district omicron variant patient profile – early features. 2021. Available from: Available from: https://www.samrc.ac.za/news/tshwane-district-omicron-variant-patient-profile-early-features (Accessed January 2022).

5. Hoffmann M, Krüger N, Schulz S, et al. The Omicron variant is highly resistant against antibody-mediated neutralization: Implications for control of the COVID-19 pandemic. Cell. 2021.

6. Abdullah F, Myers J, Basu D, et al. Decreased severity of disease during the first global omicron variant covid-19 outbreak in a large hospital in tshwane, south africa. Int J Infect Dis. 2021;116:38–42.

7. World Health Organization. Enhancing response to Omicron SARS-CoV-2 variant. 2022. Available from: https://www.who.int/publications/m/item/enhancing-readiness-for-omicron-(b.1.1.529)-technical-brief-and-priority-actions-for-member-states

8. Drain PK. Rapid Diagnostic Testing for SARS-CoV-2. N Engl J Med. 2022;386:264–72.

9. US Food and Drug Administration. SARS-CoV-2 viral mutations: Impact on Covid-19 tests. 2022. Available from: https://www.fda.gov/medical-devices/coronavirus-covid-19-and-medical-devices/sars-cov-2-viral-mutations-impact-covid-19-tests#omicronvariantimpact.

10. Rao A, Bassit L, Lin J, et al. Assessment of the Abbott BinaxNOW SARS-CoV-2 rapid antigen test against viral variants of concern. iScience. 2022:103968-.

11. Osterman A, Badell I, Basara E, et al. Impaired detection of omicron by SARS-CoV-2 rapid antigen tests. Med Microbiol Immunol. 2022:1–13.

12. Schrom J, Marquez C, Pilarowski G, et al. Comparison of SARS-CoV-2 Reverse Transcriptase Polymerase Chain Reaction and BinaxNOW Rapid Antigen Tests at a Community Site During an Omicron Surge : A Cross-Sectional Study. Ann Intern Med. 2022.

13. Arya R, Kumari S, Pandey B, et al. Structural insights into SARS-CoV-2 proteins. J Mol Biol. 2021;433:166725.

14. Mendiola-Pastrana IR, López-Ortiz E, Río de la Loza-Zamora JG, et al. SARS-CoV-2 Variants and Clinical Outcomes: A Systematic Review. Life (Basel). 2022;12:170.

15. Hodcroft E, Aksamentov I, Neher R, et al. Overview of Variants in Countries -United States (Data from GISAID). 2022. updated 25 February 2022. Available from: https://covariants.org/per-country.

16. Drain P, Sulaiman R, Hoppers M, et al. Performance of the LumiraDx Microfluidic Immunofluorescence Point-of-Care SARS-CoV-2 Antigen Test in Asymptomatic Adults and Children. Am J Clin Pathol. 2021.

17. Drain PK, Ampajwala M, Chappel C, et al. A Rapid, High-Sensitivity SARS-CoV-2 Nucleocapsid Immunoassay to Aid Diagnosis of Acute COVID-19 at the Point of Care: A Clinical Performance Study. Infect Dis Ther. 2021;10:753–61.

